# A Pilot Study Comparing Speech Characteristics in People with Parkinson’s Disease and Controls Dancing Weekly Over 5-years

**DOI:** 10.1101/2025.03.25.25324640

**Authors:** Ashkan Karimi, Narges Moein, Emily D’Alessandro, Karolina A. Bearss, Sarah Robichaud, Rachel J. Bar, Joseph F.X. DeSouza

## Abstract

**Introduction:** Parkinson’s Disease (PD) is a neurodegenerative disorder that affects motor and non-motor functions. Speech impairments, such as reduced variability in pitch (F0SD) and intensity (IntSD), are commonly observed. Early identification of these changes through voice biomarkers offers a noninvasive approach for detecting PD, tracking disease progression, and investigating the effects of interventions on this population. This study investigates the impact of dance on people with PD voice features over a five-year period and explores whether dance interventions can mitigate these impairments.

**Methods:** A longitudinal dance program involved 29 individuals with PD and 29 healthy controls. Voice recordings were collected before and after dance sessions over five years (2014-2019) and analyzed using machine learning models to extract F0SD and IntSD. Statistical analyses, including ANOVA and mixed-effect models, were performed using R studio to evaluate group differences, longitudinal changes, and the effects of dance on voice parameters.

**Results:** The analysis revealed a significant main effect of time on F0SD, indicating measurable changes over the study period. However, the interaction between group and time was not statistically significant, suggesting similar trends in both groups. While the PD group did not exhibit the expected decline in F0SD seen in previous studies, IntSD remained largely unchanged, suggesting it may be less responsive to intervention.

**Conclusion:** These findings demonstrate the potential of F0SD and IntSD as biomarkers for tracking PD progression. Dance interventions provide measurable benefits for F0SD, though further research is needed to determine optimal intervention duration and explore additional speech features such as jitter, shimmer, HNR, and CPP. Early and targeted interventions, such as combining dance with speech therapy, may enhance communication abilities and improve the quality of life for individuals with PD.

## 1. Introduction

### 1.1 Background on Parkinson’s Disease

Parkinson’s Disease (PD) is a progressive neurodegenerative disorder that affects approximately 2-3% of individuals over the age of 65 (Poewe et al., 2017) and is characterized by both motor and non-motor symptoms (Müller et al., 2013). By 2050, it is projected that 25.2 million people worldwide will be living with PD, representing a 112% increase from 2021 estimates (Su et al., 2025). The etiology of PD involves the degeneration of dopamine-producing neurons in the substantia nigra pars compacta (Lees et al., 2009), leading to a cascade of motor deficits, including tremor, rigidity, bradykinesia, and postural instability (J Jankovic, 2008). PD significantly impacts both motor and non-motor domains, leading to visible symptoms like tremor and gait disturbances as well as invisible ones such as depression and anxiety. These symptoms, combined with the stigma associated with the disease, contribute to reduced self-esteem, social isolation, and psychological distress, all of which deeply undermine the quality of life for people with PD (PwPD) (McDaniels et al., 2023). Given the progressive nature of PD, interventions must target both motor and non-motor symptoms to sustain the patient’s quality of life (QoL).

PD is notably associated with significant impairments in speech, which can impact up to 90% of patients (Smith & Caplan, 2018). These speech impairments often manifest as hypokinetic dysarthria, marked by a reduced range of pitch and intensity, and can severely hinder communication, thereby diminishing the QoL for those affected. However, speech impairments, including diminished pitch variability and vocal intensity, can appear early and often progress alongside other motor symptoms, posing significant challenges for communication. These vocal changes highlight the need for comprehensive therapeutic approaches that not only alleviate motor deficits but also address speech impairments.

Established drugs like Levodopa, dopamine agonists, anticholinergics, and Monoamine Oxidaze-B (MAO-B) inhibitors have been effective but come with significant limitations, including side effects and the inability to prevent disease progression (Kispotta et al., 2024). The side effects include but are not limited to, fibrosis, sleep attacks, impulse control disorders, depression, hypotension, and liver toxicity (Faulkner, 2014). While pharmacological treatments offer symptomatic relief, they do not halt disease progression and most ignore the non-motor PD symptoms. A recent review article by Kispotta et al. (2024), has suggested that alternative and complementary therapies for PD, including yoga, acupuncture, music therapy, and massage have demonstrated the potential to improve motor coordination, balance, and QoL by addressing both motor and non-motor symptoms. Consequently, there is an urgent need for alternative therapeutic strategies that not only manage symptoms but also potentially slow the progression of the disease (Dauer & Przedborski, 2003; Yang et al., 2022).

Recent research has demonstrated the significance of fundamental frequency (F0) variability as an early biomarker of PD (Favaro et al., 2023; Orozco-Arroyave et al., 2015; Xiu et al., 2024). A longitudinal case study demonstrated that F0 variability in speech decreases significantly during the prodromal period, several years before the clinical diagnosis of PD (Harel et al., 2004).

### 1.2. The Role of Dance in Parkinson’s Disease

Recent studies have suggested that physical and cognitive activities, such as dance, may offer neuroprotective benefits for PwPD (Bearss et al., 2017; Bearss & DeSouza, 2021; Bearss et al., 2024; Simon et al., 2024). Dance, in particular, has been shown to promote neuroplasticity in brain regions associated with motor control, such as supplementary motor area (SMA), putamen and caudate regions of the basal ganglia (Bar & DeSouza, 2016), thereby potentially mitigating the decline in motor and non-motor functions (Bearss et al., 2017; Bearss & DeSouza, 2021; Bearss et al., 2024). Specifically, dance interventions have been associated with improvements in balance, gait, and motor function, as well as enhanced mood and cognitive function in individuals with PD. The rhythmic and social aspects of dance may also contribute to these benefits by engaging multiple neural pathways and promoting overall brain health. Given the importance of speech as a noninvasive biomarker of disease onset and progression, analyzing the impact of dance on speech features could provide valuable insights into this intervention’s therapeutic potential.

### 1.3. Machine Learning in Speech Analysis

Machine learning is increasingly used in PD diagnosis, leveraging diverse data types such as handwriting (Drotár et al., 2014; Pereira et al., 2018), movement (M. Yang et al., 2009; Wahid et al., 2015; Pham & Yan, 2017), neuroimaging (Cherubini, Morelli, et al., 2014); Choi et al., 2017; Segovia et al., 2019), voice (Sakar et al., 2013; Ma et al., 2014), cerebrospinal fluid (LeWitt et al., 2013; Maass et al., 2020), cardiac scintigraphy (Nuvoli et al., 2020), serum biomarkers (Váradi et al., 2019), and optical coherence tomography (Nunes et al., 2019). It also allows combining modalities like MRI and SPECT (Cherubini, Nisticó, et al., 2014; Wang et al., 2017) to improve diagnostic precision and detect PD in early or atypical stages. The high diagnostic accuracy achieved emphasizes the potential of these techniques to introduce new biomarkers into clinical practice, enhancing precision and supporting more informed decision-making. In this context, machine learning techniques have emerged as powerful tools for analyzing complex datasets, including speech recordings. By leveraging machine learning models, it is possible to extract and analyze voice features with greater accuracy and efficiency than traditional methods.

### 1.4. Study Objectives

This study aims to investigate the differences in voice parameters, specifically the standard deviation of fundamental frequency (F0SD) and intensity (IntSD), between PwPD and healthy controls. Fundamental frequency corresponds to the perceptual correlate of pitch (Knight & Setter, 2021), while intensity relates to the loudness of the voice (Skodda, 2011). These features were chosen due to their relevance to speech impairments commonly observed in PwPD (Ma et al., 2020; Kowalska-Taczanowska et al., 2020; Holmes et al., 2000; Rusz et al., 2022; Skodda et al., 2011; Skodda et al., 2009). Additionally, the study explores whether regular dance practice can attenuate the progression of these vocal impairments over a five-year period.

## 2. Methodology

### 2.1. Study Design and Participants

This study involved a secondary analysis of voice recordings collected from PwPD and healthy controls who participated in two dance programs for PwPD from 2014 to 2019. Both programs are based on the Mark Morris Dance Group’s Dance for PD® protocol (Westheimer, 2017). Both programs were offered in Toronto, Canada. Sharing Dance Parkinson’s takes place at Canada’s National Ballet School (NBS). Dancing with Parkinson’s (DwP®) classes are offered at Trinity St. Paul’s Church. The participants self-reported their health status. The participants, consisting of 29 PwPD and 29 healthy controls, were evaluated over a five-year period (**Table 1**). The dance sessions were 1.25 hours long per week and voice recordings were taken before and after dance sessions as part of a broader study investigating the effects of dance on PD.

**Table 1.** Age and Gender Distribution of PwPD and Healthy Controls.

|  | Mean (Min-Max) Age in 2014 | Gender |
| --- | --- | --- |
| PwPD (n=29) | 68.2 (54-80) | Female:18; Male:11 |
| Healthy Controls (n=29) | 63.5 (50-75)* | Female:19; Male:10 |

\* The age of four participants was not available

### 2.2. Ethical Considerations

The original study was conducted with ethics approval from York University (certificate numbers: 2013-211 & 2017-296). All participants gave informed consent, allowing the use of their data in subsequent research. All data were anonymized to protect participant privacy, and data integrity and security were maintained throughout the analysis.

### 2.3. Data Collection

Participants were video recorded during the administration of the Movement Disorder Society’s Unified Parkinson’s Disease Rating Scale (MDS-UPDRS). The UPDRS was originally developed in the 1980s and in 2001, a new version of it, termed the Movement Disorder Society (MDS) sponsored by UPDRS was created. This scale has four parts: Part I, Nonmotor aspects of experiences of daily living (6 items assessed by interview and 7 items by self-assessment); Part II, Motor aspects of experiences of daily living (13 self-assessed items); Part III, Motor examination (18 items resulting in 33 scores by location and lateralization); and part IV, Motor complications (3 items for dyskinesia and 3 for fluctuation (Goetz et al., 2008). Specifically, participants were asked by the examiners to describe a picture drawing for 30-60 seconds. The picture is presented in **Figure 1**. The picture was chosen from the speech evaluation part of the Boston Diagnostic Aphasia Examination (BDAE) (Goodglass et al., 2001). The audio recordings of the drawing descriptions were extracted from video files using QuickTime Player (version 10.5) installed on MacOS 14 (Sonoma). The final dataset included 281 audio files, with 173 files from PwPD and 108 from healthy controls. Audio files with significant background noise or interruptions were excluded from the analysis. After that, 266 files remained, of which 135 files (84 files from PwPD and 51 files from healthy controls) were from before the dance session and 131 files were from after the dance session (with 80 files from PwPD and 51 files from healthy controls).

**Figure 1.**
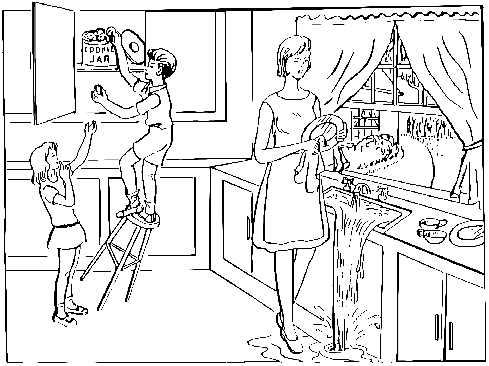
The Cookie Theft Picture (From the Boston Diagnostic Aphasia Examination – Third Edition by Harold Goodglass, Edith Kaplan, and Barbara Barresi)

Voice features were measured across 11 time points from 2014 to 2019. The 11 time points were consolidated into four to reduce variability, optimize statistical power, and address participant attrition. The decision to merge and average the 11-time points into 4 was based on both the proximity of certain data collections and the need to address variability in participant numbers across time. The first two time points, collected at the end of 2014 and the beginning of 2015, were merged due to their temporal closeness, allowing for a more accurate representation of that period. Similarly, the data from 2018 and 2019 were averaged together (time 4), as the number of voice samples in these later collections was significantly reduced due to lower participation rates. For the other two time points (2016 and 2017), an average was calculated to represent a measure for each year. Final timepoints include Time 1 (2014/2015), Time 2 (2016), Time 3 (2017), and Time 4 (2018/2019). By averaging these collections, the analysis could maintain statistical power while still capturing the longitudinal changes in voice features over the 5 years of the study. This approach provided a balanced representation across the different stages of the intervention.

### 2.4. Voice Feature Extraction

Voice features were extracted using a machine learning model implemented in Python. The primary features of interest were the standard deviation of fundamental frequency (F0SD) and intensity (IntSD), which were chosen based on their relevance to speech impairments in PD. The extraction process involved analyzing the selected voice segments for these parameters, ensuring consistency and accuracy across all recordings (**Figure 2**).

**Figure 2.**
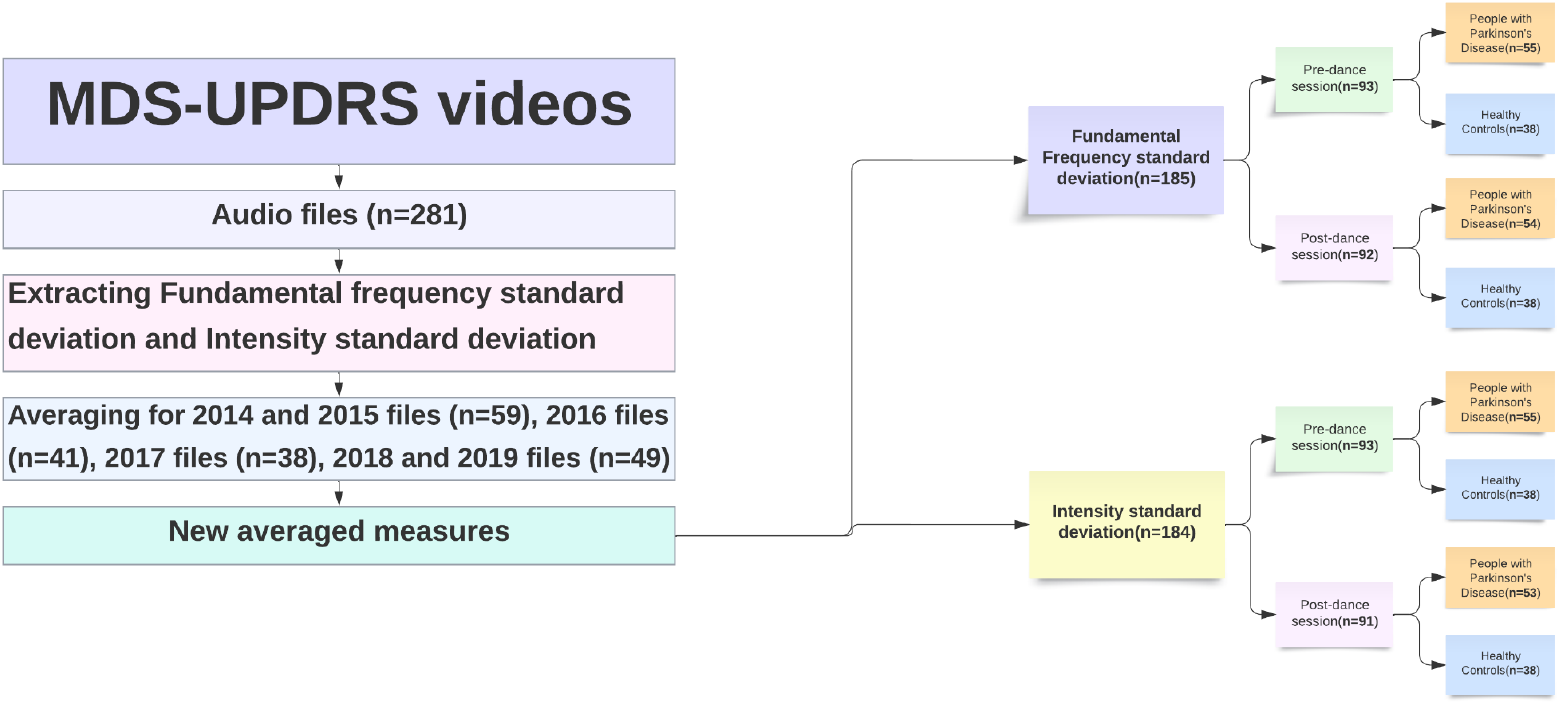
Illustration of Voice File Extraction and File Count by Group

**Figure 3.**
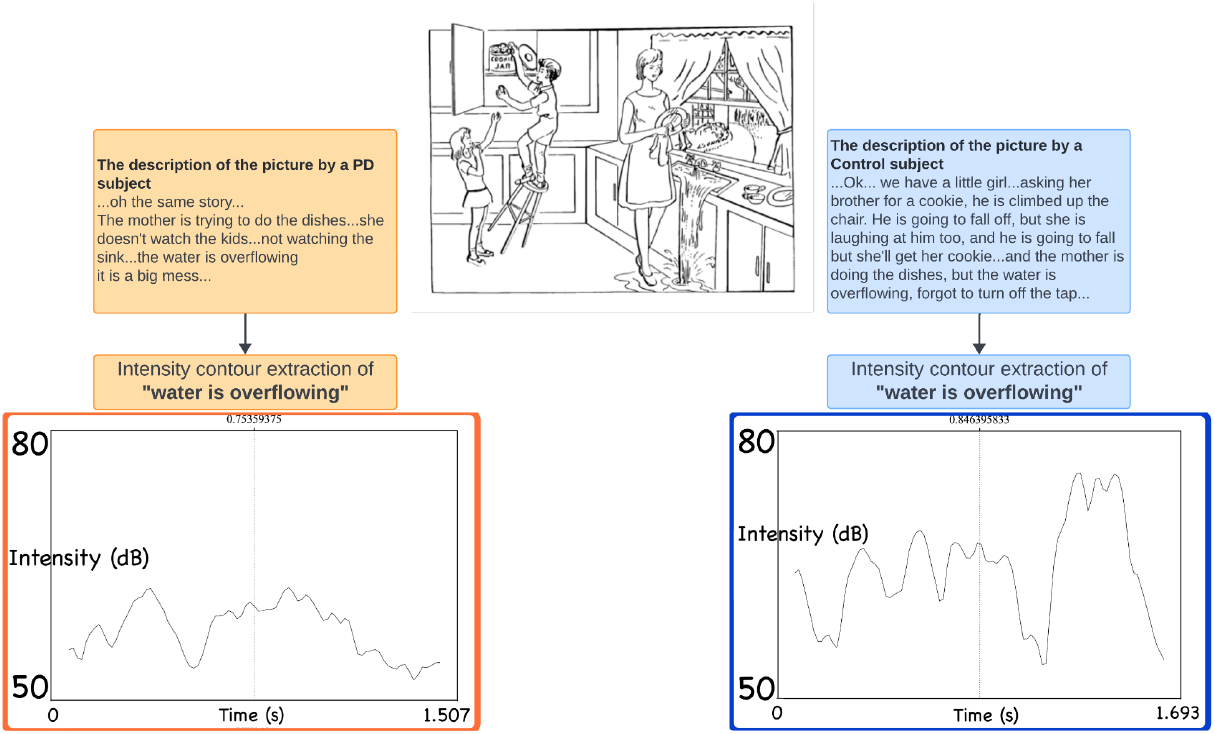
Illustration of Intensity Contours for a Common Sentence: Comparing PD and Control Subjects to Visualize Voice Intensity Differences

### 2.5. Statistical Methods

A mixed-effect ANOVA test was conducted to analyze time and group effect on F0SD and IntSD. Given that different subjects had varying numbers of samples and some had missing data for either pre- or post-dance sessions, the mixed-effect ANOVA is best suited for investigating the impact of dance on voice features in the two groups. All the statistical tests were performed in R Studio Version 2024.04.0+735.

## 3. Results

### 3.1. Effect of Dance on Voice Features

A mixed-effect ANOVA evaluated the impact of dance sessions on F0SD and IntSD. While both PwPD and healthy controls participated in pre- and post-dance evaluations, a significant main effect of time was found only for F0SD (*F* = 7.1065, *p* = 0.0084), indicating changes in F0SD measure after the dance. However, the interaction between time and group was not significant (*p* > 0.05), implying similar changes across both groups. The difference in F0SD between the two groups, PwPD and healthy controls, at the start of the study (2014), showed a trend (T-statistic: -2.0263, *p* = 0.0531), however, the comparison of 2019 for F0SD was not significant (T-statistic: -0.2321, *p* = 0.8187), indicating both groups expressed similar values to each other. For IntSD, neither the main effects nor the interactions were statistically significant, suggesting that dance did not affect IntSD variability (*p* > 0.05).

**Figures 4** and **5** illustrate the F0SD and IntSD plots for pre- and post-dance sessions from 2014 to 2019.

**Figure 4.**
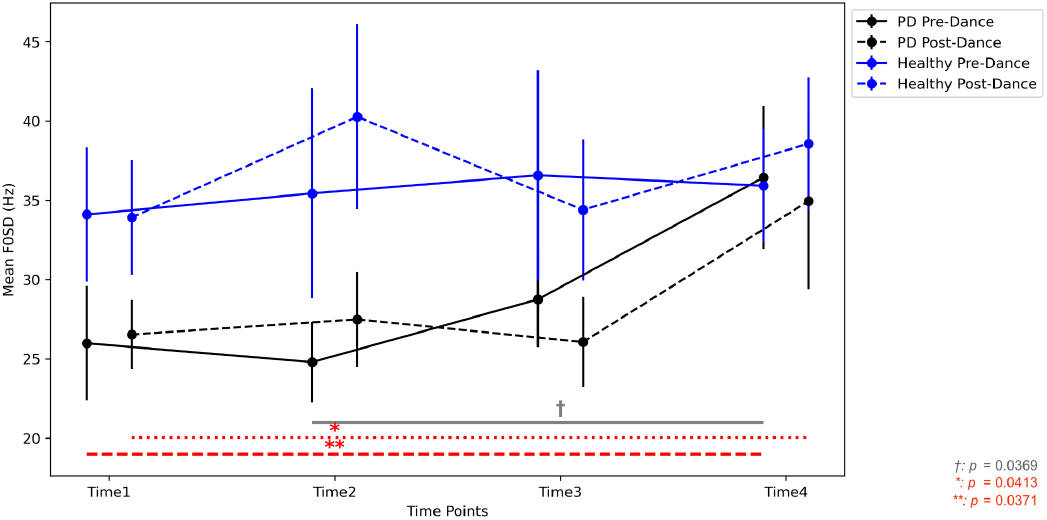
F0SD Pre- and Post-Dance Across Time Points for PwPD and Healthy Control Groups

**Figure 5.**
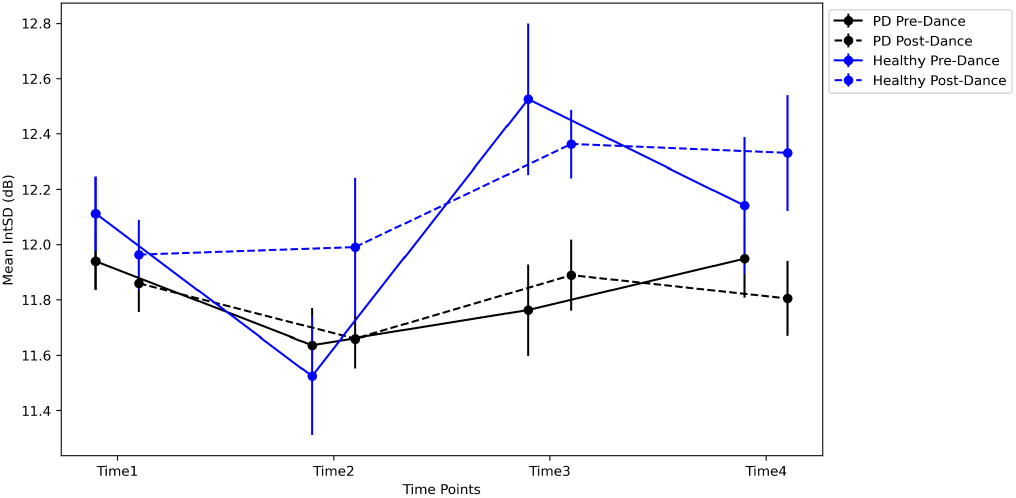
IntSD Pre- and Post-Dance Across Time Points for PwPD and Healthy Control Groups

## 4. Discussion and Conclusion

This study investigated longitudinal changes in F0SD and IntSD in PwPD and healthy controls and assessed the effects of a dance intervention over five years. Our findings contribute to the growing body of literature on speech biomarkers for PD progression (Favaro et al., 2023; Harel et al., 2004; Orozco-Arroyave et al., 2015; Xiu et al., 2024) and provide insights into how physical activity, such as dance, may influence voice features. Notably, our study showed that F0SD was significantly lower in PwPD compared to healthy controls at the start of the study, supporting its potential as a reliable speech-based biomarker for distinguishing individuals with PD. Interestingly, as participants engaged in regular dance classes, F0SD improved over time in the PD group—contrary to the expected decline associated with disease progression (Favaro et al., 2024)—highlighting the possible neuroprotective or compensatory effects of physical activity on vocal motor control.

### 4.1. F0SD as a Potential Biomarker

The results revealed a significant main effect of time on F0SD, indicating measurable changes in pitch variability over the 5-year research project. Interestingly, although longitudinal studies have reported a natural decline in F0SD with PD progression (Favaro et al., 2024), our results showed no significant reduction in F0SD within the PwPD. This unexpected preservation of pitch variability may be linked to the dance intervention, which has been suggested to enhance motor control and vocal expressiveness in individuals with PD (Bearss et al., 2017; Bearss & DeSouza, 2021; Bearss et al., 2024). One possible explanation for this preservation is the neuroplastic changes induced by the dance intervention in key motor and speech-related brain regions, particularly the supplementary motor area (SMA) and basal ganglia. The SMA plays a crucial role in motor planning and initiation, and its activation is known to be reduced in PD due to dopamine depletion (Wu & Hallett, 2013). However, studies suggest that rhythmic and repetitive movements, such as those performed in dance, may facilitate functional reorganization within the SMA, compensating for deficits in motor execution and coordination (Meulenberg et al., 2023). This reorganization could extend to vocal motor control (Dai et al., 2022; Zappa et al., 2021), explaining the maintenance of pitch variability observed in the PwPD group. Furthermore, the basal ganglia, a network heavily implicated in speech-motor control, is particularly vulnerable in PD due to its dependence on dopaminergic signaling. While progressive degeneration of these structures typically results in hypokinetic dysarthria and reduced prosodic variation (Duffy, 2012), behavioral interventions like dance have been associated with increased connectivity between the basal ganglia and cortical motor areas, potentially supporting preserved speech-motor function (Li et al., 2015). Dance-induced sensorimotor engagement helped reinforce compensatory pathways, allowing participants to retain greater control over vocal pitch variability despite disease progression. However, the interaction effect between time and group was not statistically significant, suggesting that both groups experienced similar patterns of change in F0SD over time.

These findings suggest that dance-based interventions may promote experience-dependent plasticity in motor and speech networks, enabling PwPD to sustain vocal function beyond what would be expected with natural disease progression.

### 4.2. Lack of Significant Effects on IntSD

In contrast to F0SD, no significant effects were found for IntSD, indicating that dance did not influence intensity variability in either group. This result aligns with previous research suggesting that normal aging more strongly influences vocal intensity changes rather than PD-specific mechanisms (Eichhorn et al., 2018; Ngo et al., 2022; Souza et al., 2011). The absence of significant group or time effects for IntSD suggests that intensity regulation may be less sensitive to intervention-based modulation than F0SD.

The findings indicate that IntSD alone may be an unreliable biomarker for tracking PD onset and progression. However, since previous studies have associated reduced intensity variability with PD-related dysarthria (Rusz et al., 2011), additional investigations incorporating other speech parameters (e.g., jitter, shimmer, Harmonics to Noise Ratio [HNR], Cepstral Pitch Period [CPP]) may provide a more comprehensive assessment of speech deterioration (Asgari & Shafran, 2010; Rusz et al., 2011; Upadhya et al., 2017; Vikas & Sharma, 2014).

### 4.3. Clinical and Research Implications

The preserved F0SD in PwPD suggests that dance interventions may have a protective role against speech decline in PD. To assess potential improvements in F0SD, we compared our findings with those of Favaro et al. (2024), who demonstrated that, as a natural course of PD progression, F0SD typically deteriorates over time. The contrast between our observed results and the expected decline reported by Favaro et al. (2024) provides indirect evidence that dance interventions may contribute to the preservation of F0SD (**Figure 6**). However, future studies should better incorporate non-dancing control groups to isolate the effects of dance on vocal variability.

**Figure 6.**
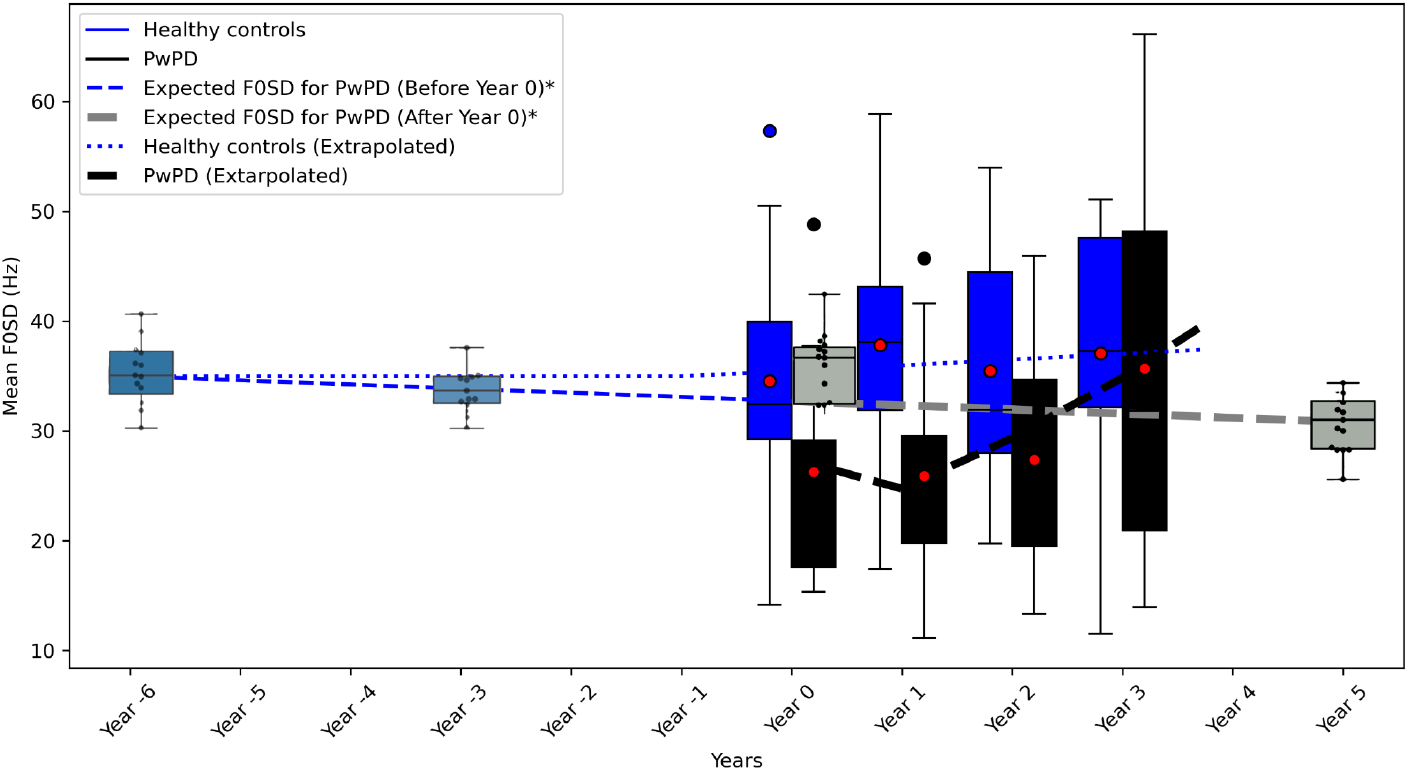
Trends in Fundamental Frequency Standard Deviation (F0SD) over Time for Healthy Controls and People with Parkinson’s Disease (PwPD). The blue boxplots represent F0SD data from healthy controls, and the black boxplots represent data from PwPD participants in this study.* The expected trendline for PwPD, based on Favaro et al. (2024), is shown in blue (before Year 0) and grey (after Year 0), indicating an expected decline in F0SD over time. In contrast, the black line illustrates the observed trend in F0SD for PwPD in this study, showing an upward trend. The preservation and slight increase in F0SD (F = 7.1065, *p* = 0.0084^*^) among PwPD may be attributed to the dance intervention over the study period.

### 4.4. Limitations

Given the exploratory nature of this research, several methodological considerations should be acknowledged. First, confounding factors may have influenced the findings, as the study did not account for the duration or stage of Parkinson’s disease in participants. This omission could have contributed to variability in F0SD trends. Additionally, the study lacked control groups for dance participation, meaning there were no comparisons with individuals with Parkinson’s disease or healthy controls who did not engage in dance sessions. As a result, it is difficult to definitively attribute the observed effects to dance alone.

Another limitation is the small sample size, which may have reduced the study’s statistical power and limited its ability to detect subtle intervention effects. Furthermore, intra-group variability may have played a role in the results, as differences in disease severity, medication status, and individual responsiveness to dance could have influenced outcomes. Lastly, given the observational study design, the absence of a randomized controlled trial (RCT) prevents any firm conclusions about causality.

To address these limitations, future research should expand the sample size to enhance statistical power and implement RCTs to better control for confounding variables. Additionally, examining a broader range of acoustic features beyond F0SD and IntSD could provide a more comprehensive understanding of speech changes. Extending the duration of the intervention or increasing session intensity would help determine the long-term effects of dance on speech function. Finally, incorporating multimodal approaches, such as combining dance with speech therapy, could offer a more effective therapeutic strategy for individuals with Parkinson’s disease.

### 4.5. Conclusion

Our findings suggest that F0SD remains a promising biomarker for tracking PD progression, with potential benefits from dance interventions. However, the lack of a significant interaction effect indicates that more rigorous studies are needed to confirm whether dance actively mitigates PD-related speech decline. The non-significant findings for IntSD highlight the need for further investigation into its role in voice-based assessments.

Ultimately, this study reinforces the importance of speech analysis as a noninvasive tool for monitoring Parkinson’s disease and points to the potential role of dance interventions in preserving vocal function. Continued investigations integrating machine learning models, speech biomarkers, and movement-based therapies could lead to more effective therapeutic strategies for people with Parkinson’s disease.

## Data Availability

All data produced in the present study are available upon reasonable request to the authors.

